# Timing of changes in Alzheimer’s disease plasma biomarkers as assessed by amyloid and tau PET clocks

**DOI:** 10.1101/2024.10.25.24316144

**Authors:** Marta Milà-Alomà, Duygu Tosun, Suzanne E. Schindler, Zachary Hausle, Yan Li, Kellen K. Petersen, Jeffrey L. Dage, Lei Du-Cuny, Ziad S. Saad, Benjamin Saef, Gallen Triana-Baltzer, David L. Raunig, Janaky Coomaraswamy, Michael Baratta, Emily A. Meyers, Yulia Mordashova, Carrie E. Rubel, Kyle Ferber, Hartmuth Kolb, Nicholas J. Ashton, Henrik Zetterberg, Erin G. Rosenbaugh, Martin Sabandal, Leslie M. Shaw, Anthony W. Bannon, William Z. Potter, Alzheimer’s Disease Neuroimaging Initiative (ADNI), Foundation for the National Institutes of Health (FNIH) Biomarkers Consortium Plasma Aβ and Phosphorylated Tau as Predictors of Amyloid and Tau Positivity in Alzheimer’s Disease Project Team

## Abstract

Plasma biomarkers for Alzheimer’s disease (AD) are increasingly being used to assist in making an etiological diagnosis for cognitively impaired (CI) individuals or to identify cognitively unimpaired (CU) individuals with AD pathology who may be eligible for prevention trials. However, a better understanding of the timing of plasma biomarker changes is needed to optimize their use in clinical and research settings. The aim of this study was to evaluate the timing of change of key AD plasma biomarkers (Aβ42/Aβ40, p-tau217, p-tau181, GFAP and NfL) from six different companies, along with established AD biomarkers, using AD progression timelines based on amyloid and tau PET.

We used data from the Alzheimer’s Disease Neuroimaging Initiative (ADNI), including 784 individuals with longitudinal ^18^F-florbetapir amyloid PET and 359 individuals with longitudinal ^18^F-flortaucipir tau PET, to estimate age at amyloid and tau positivity, defined as the age at the first positive PET scan. Of these, longitudinal plasma biomarker measures were available from 190 individuals with an estimated age at amyloid positivity and 70 individuals with an estimated age at tau positivity. Age at tau positivity was a stronger predictor of symptom onset than age at amyloid positivity in 17 individuals who progressed from CU to CI during their participation in the ADNI study (Adj R^2^ = 0.86 *vs.* Adj R^2^ = 0.38), and therefore was used to estimate symptom onset age for all individuals with an estimated age at tau positivity. Generalized additive mixed models (GAMMs) were used to model biomarker trajectories across years since amyloid positivity, tau positivity, and symptom onset, and to identify the earliest timepoint of biomarker abnormality when compared to a reference group of amyloid- and tau-negative CU individuals, as well as time periods of significant change in biomarkers.

All plasma biomarkers except NfL became abnormal prior to amyloid and tau positivity. Plasma Aβ42/Aβ40 was the first biomarker to reach abnormality consistently across timelines and plasma GFAP became abnormal early in the tau timeline. Plasma Aβ42/Aβ40 levels reached a plateau, while plasma p-tau217, p-tau181, GFAP and NfL increased throughout disease progression. Some differences in the timing of change were observed across biomarker assays.

The primary utility of plasma Aβ42/Aβ40 may lie in early identification of individuals at high risk of AD. In contrast, p-tau217, p-tau181, GFAP and NfL increase throughout the estimated timelines, supporting their potential as biomarkers for staging and monitoring disease progression.

## Introduction

Alzheimer’s disease (AD) is defined by the extracellular accumulation of amyloid plaques comprised mainly of amyloid-β (Aβ) peptide and the formation of intraneuronal neurofibrillary tangles comprised of hyperphosphorylated tau^1^. Amyloid plaques and tau tangles accumulate for many years in asymptomatic individuals during the preclinical phase of AD, but higher levels of pathology are often accompanied by neurodegeneration and inflammation culminating in the symptomatic phase which includes mild cognitive impairment (MCI) and dementia^2^. Biomarkers used to detect AD brain pathology in living individuals include Positron Emission Tomography (PET) scans that visualize the burden and distribution of amyloid and tau pathology, and measurement of proteins in the cerebrospinal fluid (CSF) or blood^3^.

Blood biomarker (BBM) tests for AD are increasingly being used in research studies, clinical trials, and clinical practice^4^. Since blood collection is more feasible and accessible than PET or CSF, BBM tests are expected to expand access to AD biomarker testing, which may improve diagnostic accuracy for patients with cognitive impairment, increase access to AD-specific treatments, and accelerate the recruitment of individuals for clinical trials^5,6^. Available BBM tests include the ratio of Aβ42 to Aβ40 (Aβ42/Aβ40)^7^, concentrations of tau phosphorylated at position 181 and 217 (p-tau181, p-tau217)^8^, the ratio of p-tau217 to non-phosphorylated tau (%p-tau217)^9^, and concentrations of neurofilament light (NfL) and glial fibrillary acidic protein (GFAP)^10,11^. BBM tests, including those for Aβ42/40 and p-tau217, start to change during the preclinical phase of AD and are used by some clinical prevention trials to identify individuals at risk of cognitive decline^12^. Additionally, some BBM levels change in response to treatments, suggesting a potential role in treatment monitoring^13,14^.

Given the potential of BBMs to accelerate development of AD treatments, the Biomarker Consortium for the Foundation for the National Institutes of Health (FNIH), which includes stakeholders from academia, industry, and patient advocacy groups, coordinated measurements of plasma samples from the Alzheimer’s Disease Neuroimaging Initiative (ADNI) with BBM tests from six different diagnostics companies. While the study focused on Aβ42/Aβ40, p-tau217, and p-tau181, panels from two companies additionally included GFAP and NfL. An initial cross-sectional study compared the classification accuracies and correlations of the BBM test measures with amyloid PET, tau PET, cortical thickness, and cognitive impairment^15^. A major additional aim of the FNIH project is to understand the timing of plasma biomarker changes, including the number of years before symptom onset certain plasma biomarkers change, which may be helpful in identifying individuals who are likely to experience cognitive decline over the next several years.

The trajectories of plasma biomarkers may vary with disease stage, which might not be fully captured when assessed as a function of chronological age, given the variability in the ages at which individuals develop AD pathology and related symptoms. Estimating timelines based on disease pathophysiology and symptomatology may provide a more accurate understanding of biomarker dynamics throughout disease progression^16–18^. One approach is to use the consistent accumulation of amyloid pathology as measured by amyloid PET as a clock, which enables calculation of the age at the start of amyloid accumulation and estimation of the age at AD symptom onset for an individual^19,20^. Biomarker values can then be aligned by years from estimated symptom onset for each individual, enabling modeling of the years before symptom onset that different biomarkers change^18^. In this study, we created clocks using longitudinal amyloid PET and tau PET data, estimated the age at amyloid positivity and tau positivity, and estimated the age at AD symptom onset for individuals based on the age at tau positivity. The trajectories of plasma biomarkers, along with established AD biomarkers, were then evaluated as a function of estimated years from amyloid positivity, tau positivity, and AD symptom onset, and the relative timing of biomarker changes were determined. Notably, all datasets used by this study, including the estimated ages of amyloid and tau PET positivity and symptom onset, are available at adni.loni.usc.edu.

## Materials and methods

### 1. Study participants

The current study utilized data from the ADNI database (adni.loni.usc.edu), which aims to develop and validate biomarkers for AD clinical trials^21,22^. ADNI was launched in 2003 as a public-private partnership led by Principal Investigator Michael W. Weiner, MD. The primary goal of ADNI has been to test whether serial magnetic resonance imaging (MRI), PET, other biological markers, and clinical and neuropsychological assessment can be combined to measure the progression of mild cognitive impairment (MCI) and early AD. All ADNI sites had local institutional approvals and written informed consent and HIPPA forms were signed by each participant, or their legally authorized representative, at their study site. Race and sex were self-identified.

Amyloid and tau PET clock models were created using longitudinal ^18^F-florbetapir (FBP) amyloid PET (*n*=784) and ^18^F-flortaucipir (FTP) tau PET (*n*=359) data regardless of the availability of plasma biomarkers.

The timing of plasma biomarker change was modeled using data from the FNIH Biomarkers Consortium “trajectories” study (*n*=292) dataset, hereafter referred to as the “main study cohort”. FNIH dataset included CU and CI ADNI participants who had plasma samples collected within six months of an amyloid PET scan for three distinct time points^15^. For individuals with more than three time points, plasma samples were selected that represented the earliest, latest, and an intermediate timepoint. For modeling the timing of change in established (non-plasma) AD biomarkers and the Clinical Dementia Rating Sum of Boxes (CDR-SB), ADNI data were not restricted to the FNIH dataset (*n*=757 for CDR-SB, *n*=743 for amyloid PET, *n*=627 for cortical thickness, *n*=569 for CSF p-tau181/Aβ42 and *n*=323 for tau PET). Positivity for either amyloid PET or tau PET at one timepoint was required for estimating age at amyloid and tau positivity, as explained below.

### 2. Plasma biomarker measurements

Plasma samples were collected, processed, and stored according to ADNI protocols^23^. As has previously been described, samples underwent analysis with C2N Diagnostics PrecivityAD2 (C2N), Fujirebio Diagnostics Lumipulse (Fujirebio), ALZpath Quanterix (ALZpath), Janssen LucentAD Quanterix (Janssen), and Roche Diagnostics NeuroToolKit (Roche) assays; a subset (84%) additionally underwent analysis with the Quanterix Neurology 4-Plex (Quanterix) assays^15^.

### 3. CSF biomarker measurements and *APOE* genotyping

CSF Aβ42, total tau, and p-tau181 were measured using the Elecsys® β-amyloid (1–42), Elecsys Total-Tau, and Elecsys Phospho-Tau (181P) immunoassays, respectively, on a Cobas e 601 analyzer (Roche Diagnostics International Ltd, Rotkreuz, Switzerland). *APOE* genotyping was performed as part of the ADNI protocol.

### 4. Amyloid PET, tau PET, and MRI measures

Amyloid PET imaging was conducted at each ADNI site following standardized protocols for FBP^24^. A global standardized uptake value ratio (SUVR) was estimated across cortical summary regions, specifically the frontal, cingulate, parietal, and lateral temporal cortices, as defined by FreeSurfer v7.1. This ratio was normalized to a composite reference region as recommended in ADNI for longitudinal analyses. The composite reference region is a non-weighted average of whole cerebellum, brainstem/pons, and subcortical WM regions proposed by Koeppe^25^. Aβ PET positivity threshold was set at 0.78 SUVR for FBP, as specified for ADNI study, which corresponds roughly to 20 Centiloid units (ADNI_UCBerkeley_AmyloidPET_Methods_v2_2023-06-29.pdf).

Tau PET imaging was conducted at each ADNI site following standardized protocols for FTP. SUVRs were estimated for regions of interest (ROIs) as defined by FreeSurfer v7.1. A mesial-temporal meta-ROI was calculated that included regions with early tau PET signal, e.g., entorhinal, parahippocampus and amygdala. SUVRs were normalized to the inferior cerebellar grey matter reference region, as defined by the SUIT template^26^. A Gaussian mixture model with two components was applied to derive a threshold for early tau PET positivity using cross-sectional mesial-temporal meta-ROI data from the entire ADNI study (*n*=907), regardless of the availability of plasma biomarkers. The threshold was established at a value equal to the mean plus two standard deviations of the first component, which corresponds to the low tau burden cluster within the Gaussian mixture model. This process rendered a threshold of 1.41 SUVR for tau PET positivity (**Supplementary Fig. 1**).

Structural brain MRI data included a 3D MP-RAGE or IR-SPGR T1-weighted 3T MRI with sagittal slices. The full cortex of each subject brain was parcellated into 41 bilateral ROIs using a volumetric Desikan-Killiany-Tourville atlas, using FreeSurfer v5.1 for ADNI-GO/2 data and v6.0 for ADNI-3 data. A composite meta-ROI cortical thickness measurement was calculated that included the entorhinal, fusiform, parahippocampal, mid-temporal, inferior-temporal, and angular gyrus^27^. To address differences in image acquisition protocol and FreeSurfer version for image processing, the meta-ROI cortical thickness measure was harmonized using the ComBat-GAM method^28^. ComBAT-GAM harmonization and adjustment for normal age effects utilized all available ADNI data for unbiased modeling. Details on the harmonization of meta-ROI cortical thickness measure are provided in **Supplementary Methods**.

### 5. Neuropsychological and clinical variables

Participants underwent clinical assessments that included a detailed interview of a collateral source, a neurological examination of the participant, the Clinical Dementia Rating® (CDR®) and the CDR-SB^29^. Participants were classified by their clinical diagnosis as Cognitively Unimpaired (CU), Mild Cognitive Impairment (MCI) or Dementia (Dementia). For MCI or Dementia cases, clinicians assessed the potential etiology (AD or non-AD) based on clinical features^30^.

### 6. Statistical analyses

#### 6.1. Estimated years from amyloid and tau positivity

All available ADNI longitudinal amyloid PET and tau PET data, regardless of availability of plasma biomarker measures, were used to create amyloid and tau PET clocks by using a modified version of a previously reported approach^19^. Briefly, rates of change in amyloid (global) or tau (mesial-temporal) SUVR over time were estimated using linear mixed effect models with random intercepts and slopes. To capture non-linearity in the associations between pathology accumulation rates and pathology burden, we fitted generalized additive model (GAM) with a cubic spline to anchor rates of change as a function of the estimated pathology burden halfway through the follow-up period (midpoint SUVR).

To identify the interval over which rates of change are consistent, clock models were restricted to the midpoint SUVR range where the variance of the GAM fitted values fell below the 90th percentile of variance for all fitted values (**Supplementary Fig. 2**). The time between 0.0001 SUVR unit increases within the resulting SUVR range (0.62 to 1.11 SUVR for amyloid PET and 0.98 to 2.04 SUVR for tau PET), was calculated by integrating the inverse of modeled rates of change. In this study, amyloid time and tau time were conceptualized as the estimated time since amyloid or tau PET positivity. Therefore, amyloid or tau time were calculated by subtracting the cumulative estimated time at the positivity threshold (global cortical amyloid PET SUVR of 0.78 or mesial-temporal meta-ROI tau PET SUVR of 1.41) from the cumulative estimated time at each SUVR value. A more detailed explanation of the methods for amyloid and tau clock estimations is provided in **Supplementary Methods**.

Age at amyloid positivity or age at tau positivity, representing the estimated ages at which an individual first reached positivity, were estimated for those with at least one positive amyloid PET or tau PET scan available, respectively. These were calculated by subtracting the amyloid or tau time corresponding to their positive scan from their age at the scan. For individuals with multiple scans, the age at amyloid or tau positivity was averaged across all positive scans.

Time estimates were validated in individuals who converted from a negative to a positive amyloid or tau PET scan throughout the ADNI study duration (PET converters). The actual conversion age was estimated by averaging the age at the last negative scan and at the first positive scan. The actual time since conversion in PET converters was calculated by subtracting the actual conversion age from the age at the scan. Correlations were examined for the estimated time and actual time since conversion, and for the estimated and actual time intervals between scans. Estimated ages at positivity were also validated in PET converters by testing their correlation with actual conversion age. Lastly, the estimated years from amyloid or tau positivity were calculated as the difference between the age at an outcome measurement and the age at amyloid or tau positivity, respectively.

#### 6.2. Estimated years from symptom onset

We identified ADNI participants who progressed from CU at their first assessment to MCI or dementia due to AD by their last assessment and had available amyloid and tau PET data (clinical progressors). Individuals were not considered as clinical progressors if they were diagnosed with a non-AD etiology, classified as CU at their last assessment, or had a negative amyloid PET at the time of clinical progression. Symptom onset age was defined as the age at the first assessment with a CDR>0 and a clinical diagnosis of MCI or dementia due to AD.

Linear regression was used to model age at symptom onset as a function of age at amyloid positivity or age at tau positivity. We also evaluated for potential effects of *APOE* ε4 carrier status and sex. To validate the model, we evaluated the correlation of the actual symptom onset age with the estimated symptom onset age in clinical progressors. Because age at amyloid positivity was not a significant independent predictor of symptom onset age when age at tau positivity was included in the model, age at tau positivity was the only predictor in the final model for symptom onset age. This model was applied to all participants with an estimated age at tau positivity. For subsequent analyses, the estimated years from symptom onset was calculated as the age at the outcome measure minus the estimated age at symptom onset.

#### 6.3. Timing of change in outcome measures relative to estimated years from amyloid positivity, tau positivity and symptom onset

Outcome measures were modeled as a function of each estimated timeline (estimated years from amyloid positivity, tau positivity, and symptom onset). Outcome measures examined included plasma biomarkers, CSF p-tau181/Aβ42, amyloid PET (global cortical SUVR), tau PET (mesial-temporal meta-ROI SUVR), cortical thickness meta-ROI, and CDR-SB. Each outcome was modeled using generalized additive mixture models (GAMM) with cubic spline basis and random intercepts as a function of each estimated timeline, separately. The time of abnormality for each outcome measure in relation to each estimated timeline was set at the first timepoint where the confidence intervals of the model estimates did not overlap with the confidence interval of the overall mean value of a reference group of amyloid and tau PET-negative CU (CDR=0 and clinical diagnosis of CU) individuals.

The median and 95% confidence intervals of the times of abnormality were calculated using 1000 bootstrapped samples, based on the 2.5% and 97.5% thresholds. To determine the periods of significant rate of change for each outcome measure, we identified time segments where the first derivative of the function deviated significantly from zero. Approximate derivatives were found using finite differencing with Taylor series expansions.

Within the reference group of amyloid and tau PET-negative CU participants, the association between each outcome measure and age, sex and *APOE* ε4 carrier status was assessed separately using linear regression model fits. For those outcome measures that showed significant associations with age, sex or *APOE* ε4 carrier status, their raw values in the entire sample were adjusted based on the derived effects in the reference group, and the timing of changes was analyzed using the corrected values as a sensitivity analysis.

As further sensitivity analyses, the timing of changes in plasma biomarkers was analyzed restricting the main study cohort to the subset of individuals with available plasma biomarker measures across all assays (*n*=233).

All statistical analyses and figures were created using R software (R version 4.2.2).

##### Data availability

Data from this study and the study methodology report may be accessed from the ADNI Laboratory of Neuro Imaging (LONI) database: adni.loni.usc.edu.

## Results

### 1. Characteristics of the study cohorts

Data from 784 ADNI participants with longitudinal FBP PET was used for amyloid PET clock modeling (mean follow-up time of 4.7 years), and data from 359 ADNI participants with available longitudinal FTP PET (mean follow-up time of 3.0 years) was used for tau PET clock modeling (**Supplementary Table 1**).

Of the 406 participants in the main study cohort, who underwent plasma biomarker measurements as part of the FNIH project, 190 met the inclusion criteria for estimating age at amyloid positivity (i.e., had at least one positive amyloid PET scan during their participation in the ADNI study), and 70 participants satisfied the inclusion criteria for estimating age at tau positivity (i.e., had at least one positive tau PET scan during their participation in the ADNI study). 48 participants overlapped between the two cohorts. The reference group included 80 individuals who were amyloid and tau PET-negative and CU (CDR=0 and clinical diagnosis of CU) throughout the study duration and was used to establish normative distributions for AD biomarkers and cognitive measures.

The characteristics of individuals in the main study cohort with an estimated age at amyloid positivity, tau positivity, and the reference group are described at the baseline plasma sample collection (**Table 1**). In the combined cohort, mean age was 73.9 ± 7.0 years, 147 (50.3%) were women, 121 (41.4%) were *APOE* ε4 carriers, 276 (94.5%) self-identified as white, 112 (38.4%) were cognitively impaired (CDR>0) and 130 (44.5%) were amyloid PET-positive at baseline. When compared to the reference group, individuals with an estimated age at tau positivity were significantly older, while the age difference was not significant in the cohort with an estimated age at amyloid positivity. Individuals with an estimated age at amyloid or at tau positivity were more likely to carry an *APOE* ε4 allele, and those with an estimated age at amyloid positivity had significantly lower education levels than the reference group. All AD biomarkers were significantly elevated at baseline in both cohorts compared to the reference group. Notably, only 8 participants in the cohort with an estimated age at amyloid positivity and 3 participants in the cohort with an estimated age at tau positivity had a clinical diagnosis of dementia at baseline.

**Table 1.**
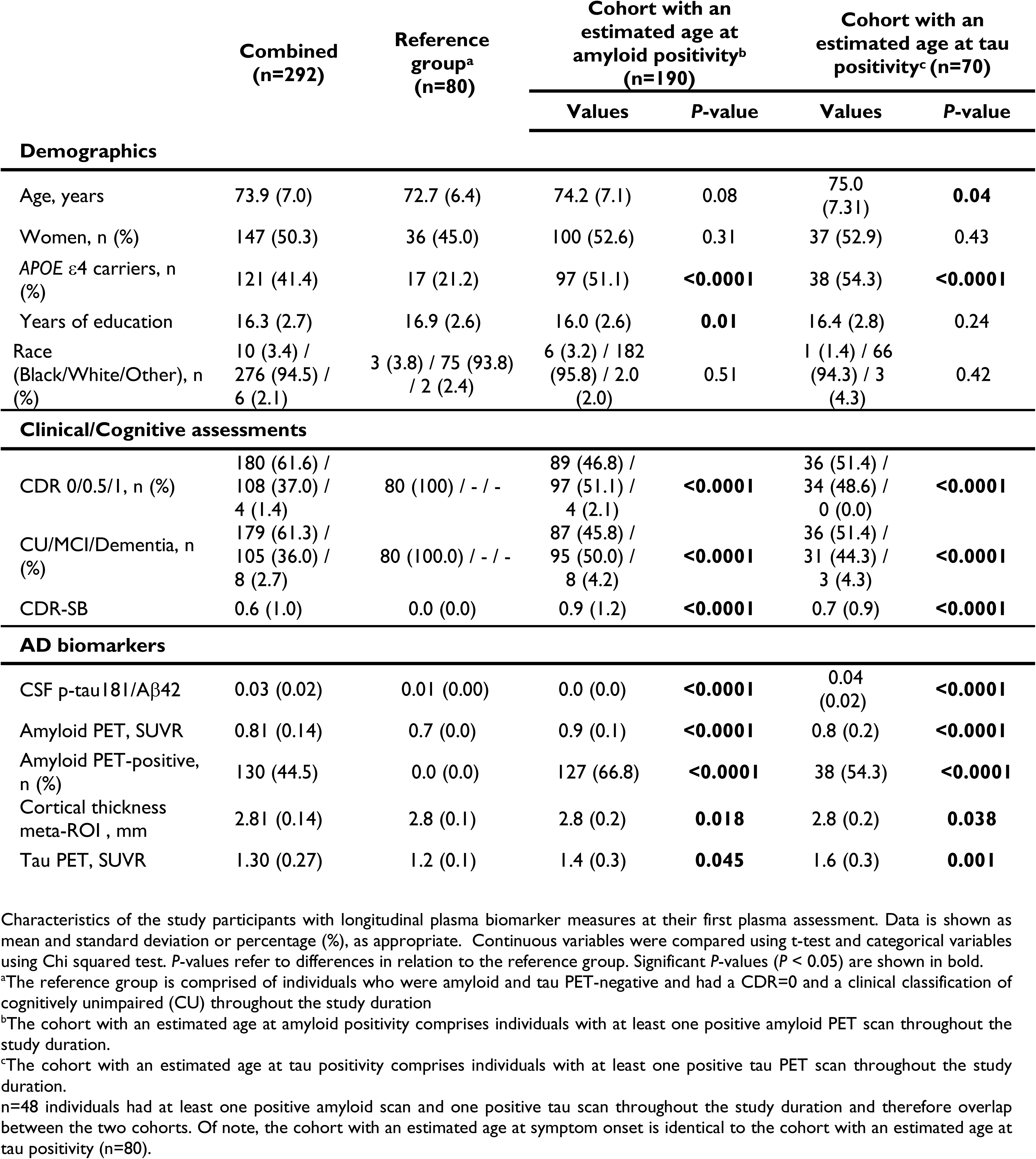
Characteristics of participants in the main study cohort.

Baseline characteristics of individuals included in the analyses for established (non-plasma) AD biomarkers and CDR-SB in the cohorts with an estimated age at amyloid (n=635) and tau (n=212) positivity and in the reference group (n=263), are described in **Supplementary Table 2**. Of note, in this larger sample, individuals in both cohorts were significantly older than those in the reference group.

Finally, among all individuals with both an estimated age at amyloid and tau positivity, 17 progressed from CU to MCI or dementia due to AD during their participation in the ADNI study and were included in the model used to estimate symptom onset age.

### 2. Creation of clocks with longitudinal ADNI amyloid PET and tau PET data

Clocks relating amyloid or tau PET SUVR to time were created and validated within the full ADNI study dataset before being applied to the main study cohort. A detailed description of the method and model validations can be found in **Supplementary Methods**.

The estimated ages at amyloid and tau positivity were highly correlated with the actual age at conversion among individuals who converted from a negative to a positive PET scan. For amyloid PET converters (n=81), the relationship was: ‘age at amyloid positivity = 1.01 x (conversion age) – 2.39 years ± 2.47 years’; Intercept *P* = 0.34; Spearman’s ρ = 0.96, *P <* 0.0001. For tau PET converters (n=35), the relationship was: ‘age at tau positivity = 1.07 x (conversion age) – 6.68 years ± 3.33 years’; Intercept *P* = 0.05; Spearman’s ρ = 0.96, *P <* 0.0001; **Supplementary Fig. 3**).

In the main study cohort, the estimated ages at amyloid positivity, tau positivity, and symptom onset were 66.6 ± 10.2, 73.7 ± 9.5, and 81.0 ± 6.1 years (mean ± standard deviation), respectively.

The observed amyloid PET SUVR or tau PET SUVR values by age are displayed in **Fig. 1A** and **Fig. 1C**, respectively, demonstrating wide variability in the ages at which participants start to accumulate amyloid and tau pathology. **Fig. 1B** and **Fig. 1D** show the modeled biomarker patterns as a function of estimated time from amyloid positivity and tau positivity, respectively, and demonstrate the relatively consistent accumulation of these pathologies over time.

**Figure 1.**
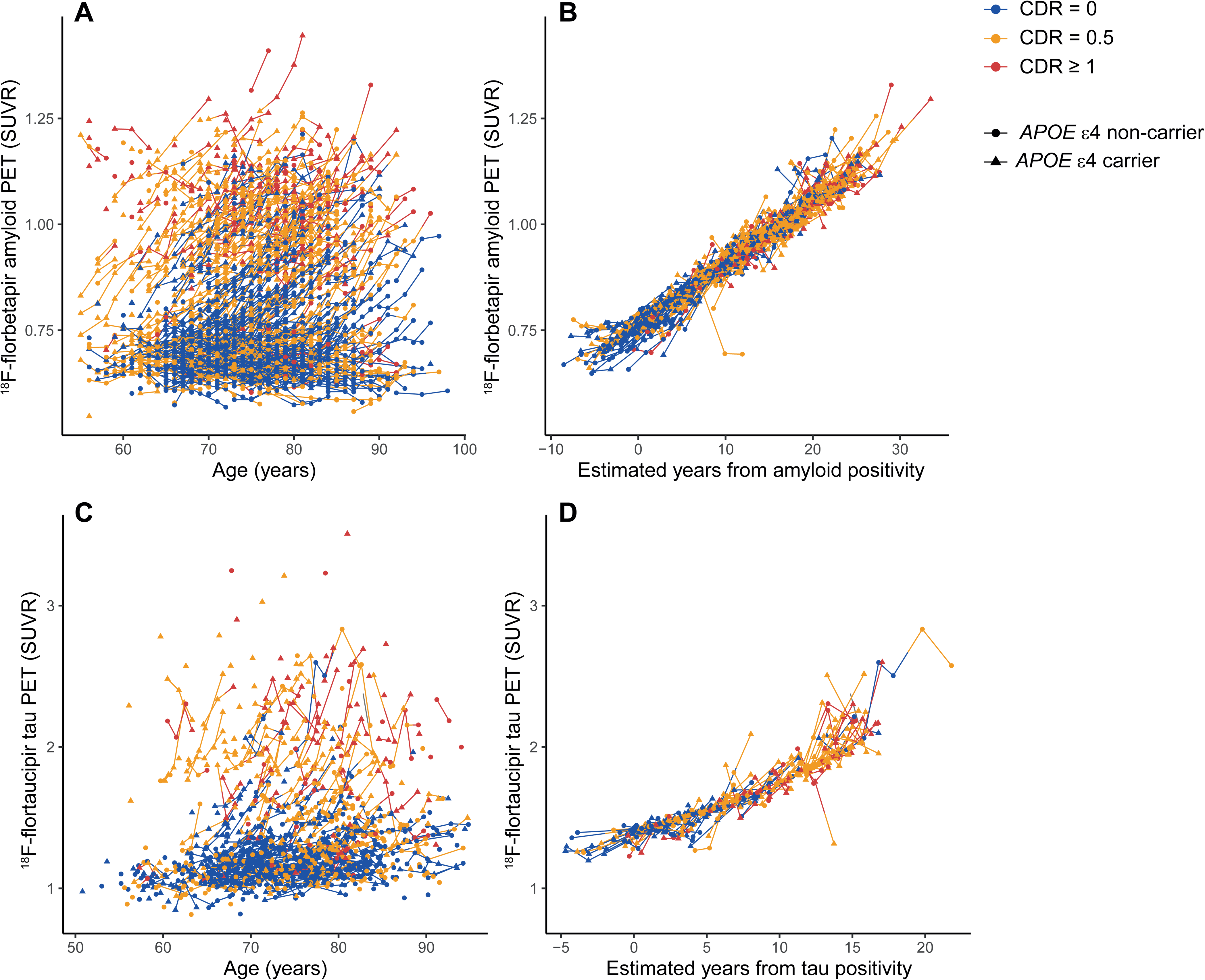
Amyloid and tau PET trajectories as a function of age and estimated years from amyloid or tau positivity. Individual amyloid (**A, B**) and tau (**C, D**) PET longitudinal trajectories, where each line connects multiple observations from the same individual. Points and line segments are color-coded according to the cognitive status assigned to each observation (Blue for CDR=0, orange for CDR=0.5, and red for CDR≥1). Triangles represent *APOE* ε4 carriers and circles represent *APOE* ε4 non-carriers. One value with a ^18^F-flortaucipir tau PET SUVR>4 was excluded for visualization.

### 3. Estimation of age at symptom onset

Using the full ADNI dataset, the age at amyloid positivity for the clinical progressors (*n*=39) was predictive of the age at symptom onset (Spearman’s ρ=0.63; Adj R^2^=0.38; *P*<0.0001). However, the age at tau positivity was a much stronger predictor of the age at symptom onset (*n*=17; Spearman’s ρ=0.93; Adj R^2^=0.86; *P* < 0.0001; **Supplementary Fig. 4**). When both age at amyloid and tau positivity were included in the model, age at amyloid positivity did not show a significant effect (*P* > 0.05). Importantly, neither *APOE ε4* status nor sex significantly influenced the estimated age at symptom onset in these models. Therefore, age at tau positivity was included as the only predictor in the final model used to estimate symptom onset. A detailed description of the method and model validations can be found in **Supplementary Methods.**

### 4. AD biomarker trajectories as a function of years from amyloid positivity, tau positivity and symptom onset

Trajectories of AD plasma biomarkers, established AD biomarkers, and CDR-SB are shown as a function of the three estimated AD progression timelines in **Fig. 2** and **Fig. 3**. Plasma biomarkers previously reported to be most strongly associated with amyloid PET and tau PET are shown in **Fig. 2**^15^; trajectories for the remaining plasma biomarkers are provided in **Supplementary Fig. 5** and **Supplementary Fig. 6**. Trajectories of all outcome measures as a function of age, amyloid PET, and tau PET are depicted in **Supplementary Figs. 7-9**.

**Figure 2.**
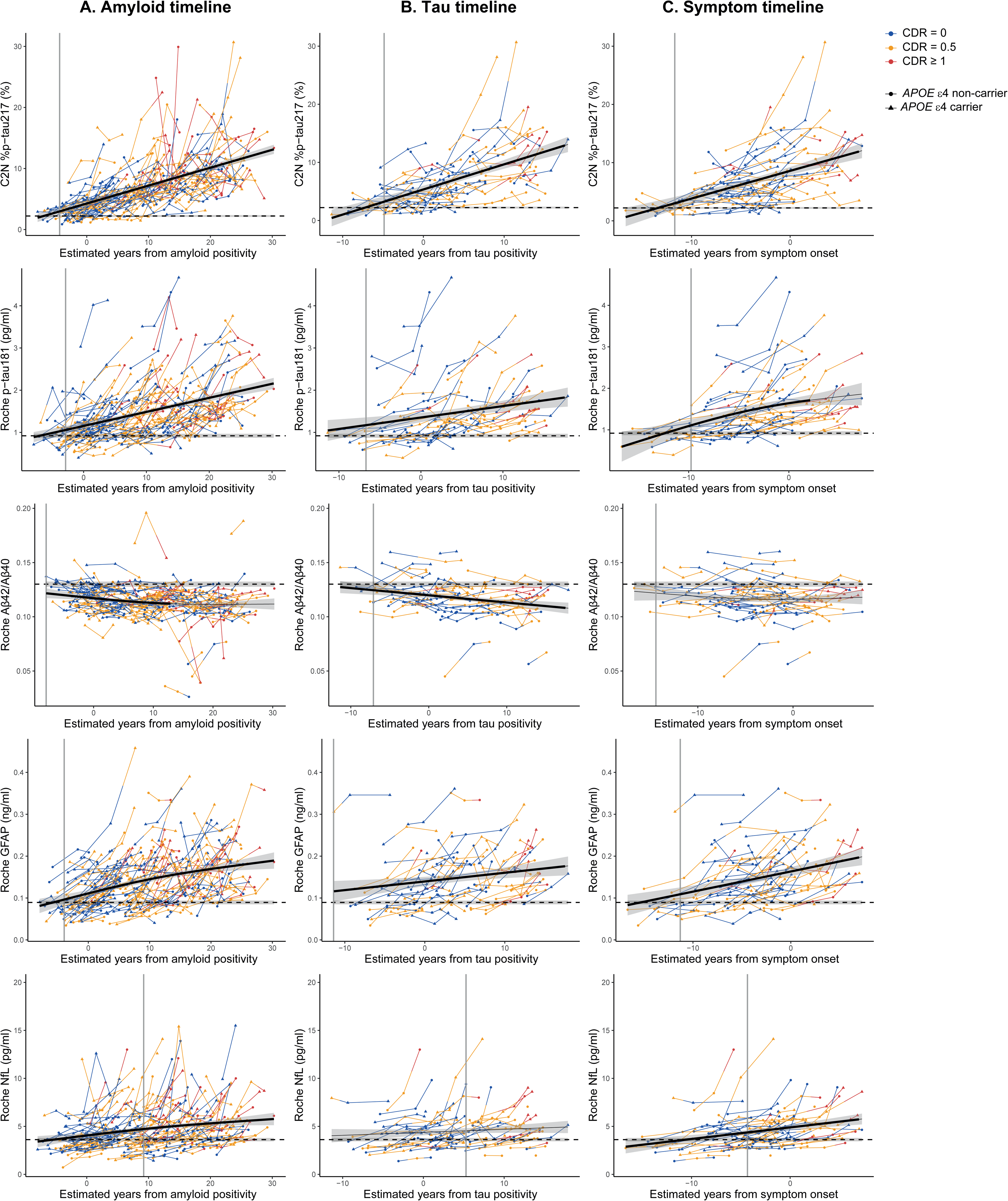
AD plasma biomarkers as a function of AD progression timelines. Generalized Additive Mixed Models (GAMM) with cubic spline basis and random intercepts were used to model plasma biomarkers as a function of years from amyloid positivity **(A)**, years from tau positivity **(B)** and years from symptom onset **(C).** Dashed horizontal lines indicate the mean of the reference group, with shaded areas representing the 95% CI. Solid vertical lines mark the time of abnormality compared to the reference group. Time periods with a significant rate of change are indicated with thicker lines. Individual trajectories are shown, where each line connects multiple observations from the same individual. Points and line segments are color-coded according to the cognitive status assigned to each observation (Blue for CDR=0, orange for CDR=0.5, and red for CDR≥1). Triangles represent *APOE* ε4 carriers and circles represent *APOE* ε4 non-carriers. Outlier values were excluded for visualization (1 for Roche Ab42/Ab40, 2 for Fujirebio Ab42/Ab40, 1 for Janssen p-tau217, 2 for C2N p-tau217, 2 for ALZpath p-tau217, 3 for Fujirebio p-tau217, 3 for Quanterix p-tau181, 3 for Roche GFAP, 2 for Quanterix GFAP, 1 for Roche NfL and 1 for Quanterix NfL).

**Figure 3.**
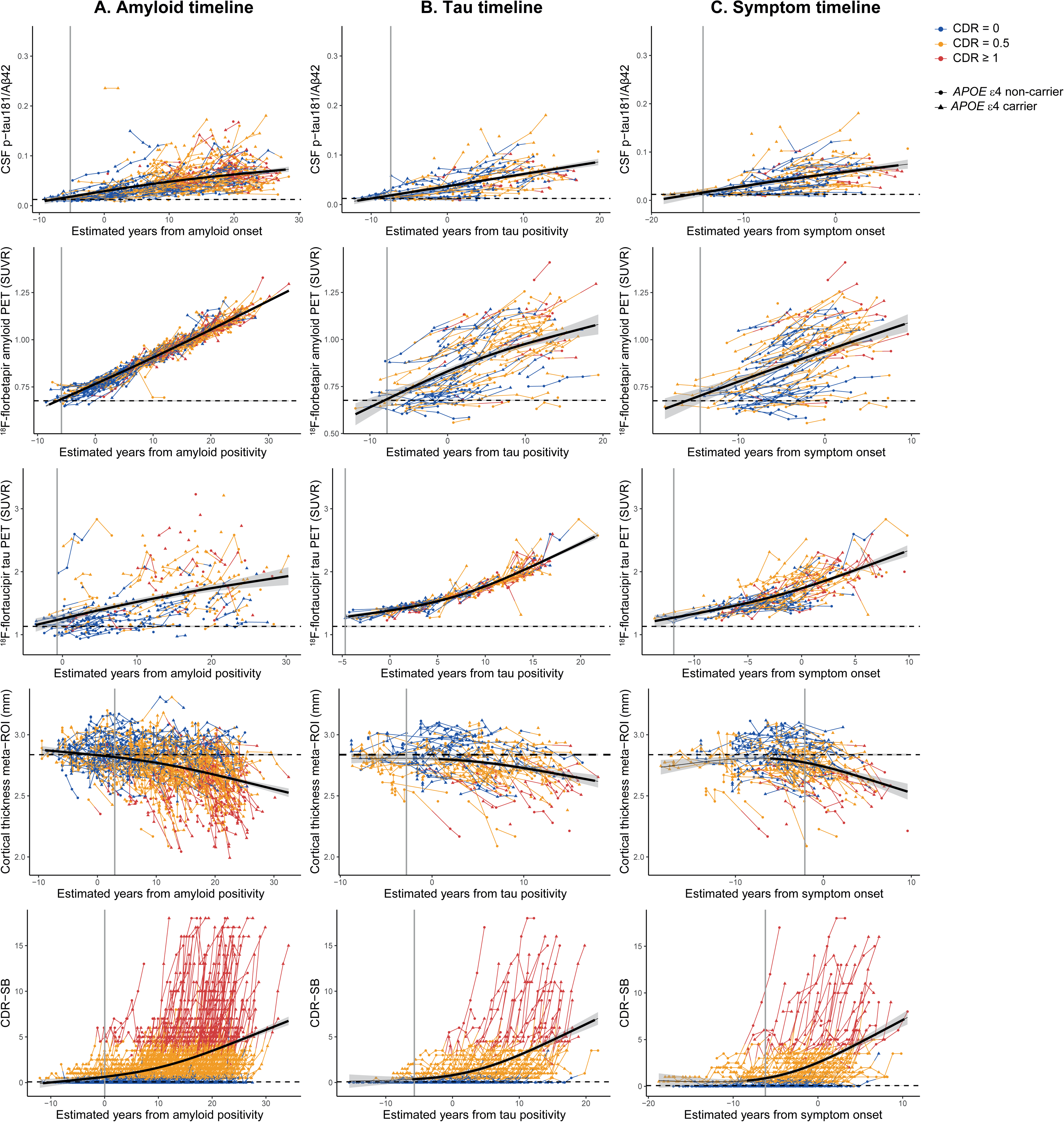
Established AD biomarkers and CDR-SB as a function of AD progression timelines. Generalized Additive Mixed Models (GAMM) with cubic spline basis and random intercepts were used to model established AD biomarkers and CDR-SB as a function of years from amyloid positivity **(A)**, years from tau positivity **(B)** and years from symptom onset **(C).** Dashed horizontal lines indicate the mean of the reference group, with shaded areas representing the 95% CI. Solid vertical lines mark the time of abnormality compared to the reference group. Time periods with a significant rate of change are indicated with thicker lines. Individual trajectories are shown, where each line connects multiple observations from the same individual. Points and line segments are color-coded according to the cognitive status assigned to each observation (Blue for CDR=0, orange for CDR=0.5, and red for CDR≥1). Triangles represent *APOE* ε4 carriers and circles represent *APOE* ε4 non-carriers. 1 value with ^18^F-flortaucipir tau PET SUVR>4 was excluded for visualization.

### 5. Timing of AD biomarker changes relative to estimated years from amyloid positivity

We modeled the longitudinal trajectories of all outcome measures as a function of estimated years from amyloid positivity and estimated the time when each measure differed significantly from the reference group mean.

All plasma biomarkers, except for NfL, reached abnormal levels before the estimated time of amyloid positivity, with plasma Aβ42/Aβ40 measures becoming abnormal first **(Fig. 4A, Supplementary Table 3)**. Specifically, C2N Aβ42/Aβ40 and Roche Aβ42/Aβ40 showed abnormal levels at a median of -7.9 years before amyloid positivity, earlier than amyloid PET (-5.9 years) or CSF p-tau181/Aβ42 (-5.2 years). Notably, Fujirebio and Quanterix Aβ42/Aβ40 measures became abnormal considerably later (-5.8 and -3.4 years, respectively). C2N %p-tau217 showed abnormality -4.4 years before amyloid positivity and was closely followed by plasma GFAP measures, Janssen and ALZpath p-tau217, Roche p-tau181 (all at median times within -3.9 to -2.9 years prior to amyloid positivity). Fujirebio p-tau217 showed abnormal levels at -1.5 years, and Quanterix p-tau181 and C2N p-tau217 did not reach abnormality until around the estimated time of amyloid positivity, along with tau PET and CDR-SB (all from - 0.9 to 0.0 years before amyloid positivity). Cortical thickness reached abnormality 2.9 years after amyloid positivity, and plasma NfL was the last to show significant elevations, 9.2 or 10.5 years after amyloid positivity, for Roche or Quanterix assays, respectively.

**Figure 4.**
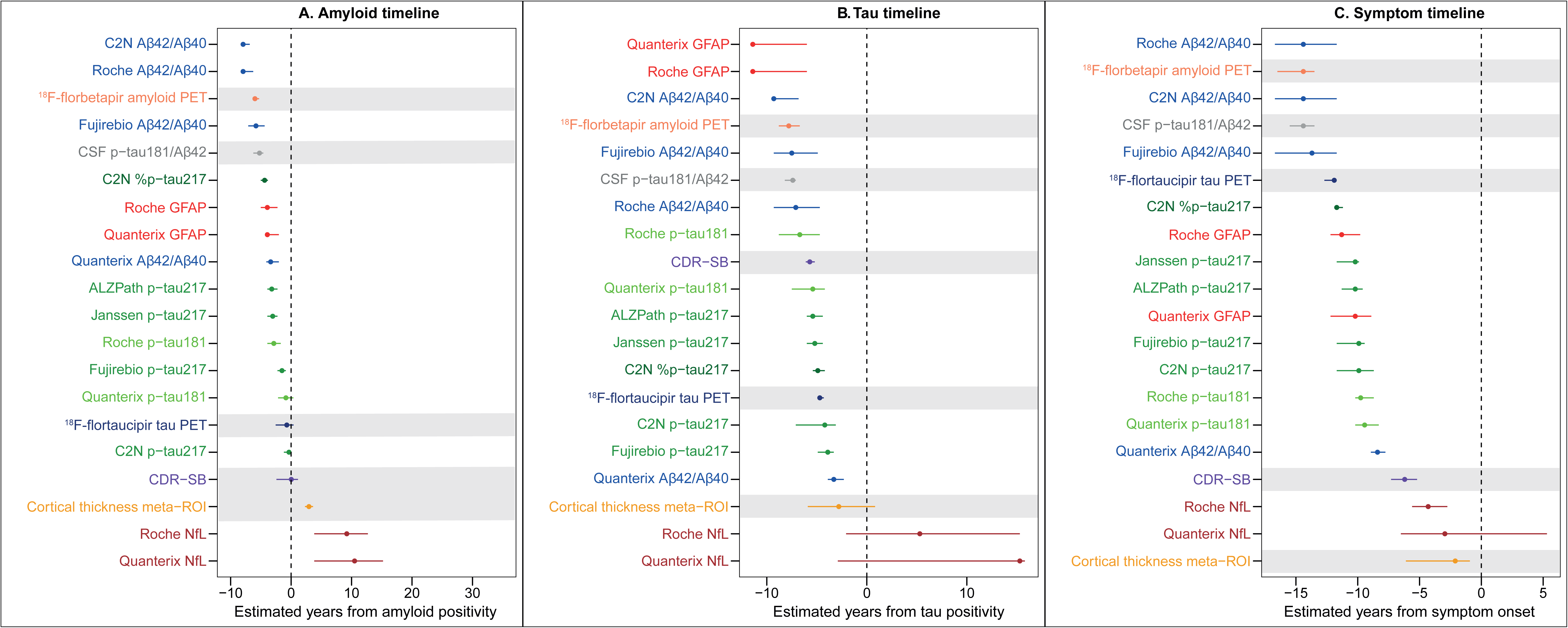
Timing for AD biomarker abnormality. Estimated years from amyloid positivity **(A),** tau positivity **(B)** or symptom onset **(C)** where each outcome measure becomes abnormal compared to the reference group. Points depict the median and error bars depict the 95% CI. Dashed vertical lines at time = 0 represent the time at amyloid positivity (global cortical ^18^F-florbetapir amyloid PET SUVR > 0.78) **(A)**, tau positivity (mesial-temporal ^18^F-flortaucipir tau PET SUVR > 1.41) **(B)** or symptom onset **(C)**. Established AD biomarkers and CDR-SB are shaded in grey.

Next, we assessed the rates of change in plasma biomarkers relative to the amyloid timeline. In **Fig.2** and **Fig.3**, thicker segments of the fitted lines indicate time periods where significant slopes were observed. All measures of p-tau217, p-tau181, GFAP and NfL consistently increased during the entire amyloid timeline ranging from -7.9 to -6.8 years before to 30.3 years after the estimated time of amyloid positivity. In contrast, plasma Aβ42/Aβ40 measures decreased early in the timeline (-7.9 to -7.1 years) but plateaued at 11.5 to 13.0 years after amyloid positivity, depending on the assay used (**Fig.2A, Supplementary Fig. 5A and 6A, Supplementary Table 4**). CSF p-tau181/Aβ42, amyloid PET, tau PET, cortical thickness and CDR-SB exhibited significant change throughout the entire amyloid timeline (**Fig. 3A, Supplementary Table 4**).

### 6. Timing of AD biomarker changes relative to estimated years from tau positivity

As a function of the tau timeline, plasma GFAP measures were the earliest to differ significantly from the reference group (up to a median of -11.5 years before estimated time of tau positivity), and were followed by C2N, Fujirebio and Roche Aβ42/Aβ40 measures (ranging from -9.3 to -7.1 years) and p-tau181 (-6.7 or -5.4 years for Roche and Quanterix, respectively). Amyloid PET and CSF p-tau181/Aβ42 also became abnormal within this time range (-7.8 and -7.4 years prior to tau positivity, respectively). Notably, Quanterix Aβ42/Aβ40 changed later than the rest of plasma Aβ42/Aβ40 measures, -3.3 years before tau positivity. Plasma p-tau217 measures became abnormal ranging from -5.4 to -3.9 years before tau positivity, and tau PET abnormalities were detected at -4.7 years. These were followed by abnormalities in cortical thickness (-2.8 years) and eventually plasma NfL (5.3 or 15.3 years after tau positivity for Roche and Quanterix, respectively). Of note, CDR-SB showed abnormality at median time of -5.7 years before tau positivity (**Fig. 4B, Supplementary Table 3**).

The temporal dynamics of plasma biomarkers across the tau timeline varied depending on the assay. C2N and Roche Aβ42/Aβ40 decreased significantly during the entire tau timeline (-11.4 years before to 17.9 years after tau positivity). In contrast, Quanterix Aβ42/Aβ40 decreased from -11.4 but then plateaued 6.0 years after tau positivity, while Fujirebio Aβ42/Aβ40 decreased significantly only from -0.8 to 4.4 years. All p-tau217 measures increased significantly throughout the entire tau timeline except for C2N p-tau217, which started to significantly rise one year before tau positivity. The increases in Roche GFAP and in Roche p-tau181 were consistent the entire tau timeline, whereas Quanterix p-tau181 rise was significant until 3.6 years after tau positivity. In contrast, Quanterix plasma GFAP and NfL measures did not significantly change at any point in the tau timeline. (**Fig. 2B, Supplementary Fig. 5B and 6B, Supplementary Table 4**).

CSF p-tau181/Aβ42, amyloid PET and tau PET consistently increased during the entire tau timeline (from -12.2 or -11.9 to 19.8 or 19.4 years for CSF p-tau181/Aβ42 and amyloid PET, respectively, and from -4.7 to 21.8 for tau PET). In contrast, CDR-SB started to change at -6.3 years prior to tau positivity. Finally, cortical thickness started to decrease shortly after tau positivity (0.5 years) and persisting until the timeline upper limit (18.1 years; **Fig. 3B, Supplementary Table 4**)

### 7. Timing of AD biomarker changes relative to estimated years from symptom onset

Roche, C2N, and Fujirebio plasma Aβ42/Aβ40 were the earliest plasma biomarkers to become abnormal in the symptom timeline, concurrent with amyloid PET and CSF p-tau181/Aβ42 (up to -14.4 years before symptom onset). In contrast, Quanterix plasma Aβ42/Aβ40 did not significantly differ from the reference group until -8.4 years prior to symptom onset. Tau PET, plasma GFAP, p-tau217 and p-tau181 measures all reached abnormality ranging from -11.9 to -9.4 years from symptom onset. These were followed by abnormalities in CDR-SB (-6.2 years), and lastly, plasma NfL and cortical thickness (ranging from -4.3 to -2.1 years before symptom onset) (**Fig. 4C, Supplementary Table 3**).

Importantly, plasma Aβ42/Aβ40 measures did not have a significant rate of change after the estimated symptom onset. Fujirebio and Quanterix plasma Aβ42/Aβ40 significantly decreased from the lower limit of the symptom timeline (-16.7 years) but stabilized -3.9 and - 0.8 years prior to symptom onset, respectively, while changes in C2N plasma Aβ42/Aβ40 were significant only in the period between -11.1 to -2.2 years. The rate of change in Roche plasma Aβ42/Aβ40 was not significant at any point as a function of the symptom timeline.

In contrast, all plasma p-tau217, as well as Quanterix p-tau181, GFAP and NfL measures steadily increased across the entire symptom timeline (-16.7 to 7.3 years from symptom onset). The rise in Roche plasma p-tau181 was significant from the timeline lower limit but stabilized 2.2 years after symptom onset (**Fig. 2C, Supplementary Fig. 5C; Supplementary Fig. 6C Supplementary Table 4**).

CSF p-tau181/Aβ42, amyloid PET, and tau PET changed consistently during the entire symptom timeline (-18.7 to 7.8 years for CSF p-tau181/Aβ42, -19.5 to 9.6 years for amyloid PET, and -13.7 to 9.8 years for tau PET). CDR-SB started to change significantly at -8.6 years and persisted throughout the timeline (10.6 years after symptom onset). Finally, the rate of change in cortical thickness became significant shortly after (-6.2 years), also persisting across the symptom timeline (9.6 years after symptom onset) (**Fig. 3C, Supplementary Table 4**).

### 8. Sensitivity analyses

Sex or *APOE* ε4 carrier status did not have a significant effect in any of the outcome measures in the reference group (all *P* > 0.05). In contrast, tau PET and all p-tau181, GFAP, NfL, and p-tau217 measures, except for C2N p-tau217 and %p-tau217, were significantly associated with age. For these biomarkers, the same analyses were performed using age-corrected values. The ordering of plasma biomarkers relative to the different timelines was very similar than that of non-corrected data (**Supplementary Table 5**). Most notable differences were for plasma NfL, which became abnormal significantly earlier after age adjustment (0.1 or 4.1 years in the amyloid timeline, -5.4 or -3.3 years in the tau timeline, and at -9.6 or -8.7 in the symptom timeline for Quanterix and Roche measures, respectively).

Analyses on the dynamics of age-adjusted plasma biomarkers across the three timelines rendered very similar results to those of non-corrected biomarkers (**Supplementary Table 6**).

Notably, Fujirebio plasma p-tau217 did not significantly rise until -3.0 years before tau positivity, and the rate of change in Quanterix plasma p-tau181 did not reach statistical significance at any point in the tau timeline after age adjustment.

Finally, the timing of plasma biomarker changes was assessed in the subset of individuals with available plasma biomarkers across all assays (*n*=149 with an age at amyloid positivity, *n*=49 with an age at tau positivity, *n*=66 in the reference group), and results were very similar to those in the full main study cohort (**Supplementary Table 7 and Supplementary Table 8**).

## Discussion

The aim of this study was to evaluate the timing of plasma biomarker changes relative to AD progression timelines based on amyloid and tau PET clocks. We estimated the age at amyloid positivity and tau positivity for ADNI participants using their longitudinal PET imaging data and found that age at tau positivity was a stronger predictor of the age at symptom onset than the age at amyloid positivity. Our main findings were 1) all plasma biomarkers except NfL exhibited changes prior to the estimated time of amyloid and tau positivity, with plasma Aβ42/Aβ40 and GFAP showing the earliest alterations; 2) plasma p-tau217, p-tau181, GFAP and NfL increased consistently over the estimated AD progression timelines, while plasma Aβ42/Aβ40 only changed early in the disease; 3) the timing of plasma biomarker changes varied depending on the AD progression timeline and the assay used. These findings contribute to improving our understanding of AD pathophysiology and the dynamics of key plasma biomarkers throughout AD progression, which is crucial for optimizing their use in clinical trials and clinical practice.

Plasma Aβ42/Aβ40 biomarkers reached abnormality as early as 7.9 years before amyloid positivity, 9.3 years before tau positivity and 14.4 years before symptom onset. These were earlier or very close times to those when CSF p-tau181/Aβ42 and amyloid PET became abnormal. Our findings are consistent with studies showing changes in plasma Aβ42/Aβ40 prior to detectable amyloid accumulation in PET^31–34^, and with plasma biomarkers reflecting soluble amyloid forms prior to fibrillar amyloid detectable with amyloid PET. Plasma GFAP changed shortly after Aβ42/Aβ40 in the amyloid and symptom timelines, concurrently with p-tau217 and p-tau181 measures. Interestingly, plasma GFAP showed the earliest median time of abnormality only in the tau timeline, although it was not significantly earlier than most plasma Aβ42/Aβ40 measures, as indicated by the overlapping confidence intervals. GFAP in plasma is reported to increase as an early response to amyloid accumulation^11,35^, which may explain why it reaches abnormality close to time of amyloid biomarkers. In addition, astrocyte reactivity has been shown to modulate the association of amyloid with early tau pathology^36^. As the years from tau positivity were estimated based on a mesial-temporal meta-ROI, which included areas of early tau accumulation, the early increase of plasma GFAP in the tau timeline could potentially reflect astrocyte reactivity in individuals with amyloid pathology and within the tau accumulation pathway. Plasma p-tau181 measures also changed earlier in the tau timeline, at similar times to Aβ42/Aβ40 and earlier than p-tau217. These differences, although subtle, might be reflecting disparities between p-tau isotopes and the pathophysiology they potentially reflect^37^. Of interest, tau PET became abnormal close to the time of amyloid positivity, shortly before the threshold used for tau positivity, but more than 11 years prior to symptom onset. This demonstrates the strong association of tau accumulation with AD symptoms. Finally, cortical thickness and plasma NfL were the last to change in the three timelines, as expected for markers indicating downstream neurodegeneration. Overall, the ordering of biomarker changes was in concordance with the A/T_1_/T_2_/(N) conceptualization, in terms of amyloid biomarkers changing first and being followed by biofluid mid-region tau fragments that may reflect a response to amyloid pathology, and later on by tau PET and neurodegeneration markers^1^.

When we studied the rates of change in plasma biomarkers throughout progression timelines, decreases in plasma Aβ42/Aβ40 were subtle and plateaued at 11.5 to 13.0 years after amyloid positivity. In contrast, plasma p-tau217 and p-tau181 and, to a lesser extent, GFAP and NfL, steadily increased across AD progression similarly to established AD biomarkers, reinforcing their potential for monitoring disease progression. Plasma NfL did not significantly change in the tau timeline. As a non-specific neurodegeneration biomarker, its increase may be more evident at later disease stages. Also, multiple factors, including comorbid pathologies, may contribute to plasma NfL levels and longitudinal trajectories^38–40^.

Generally, results in the timing of plasma biomarker changes were consistent across assays, but some differences were observed. Quanterix Aβ42/Aβ40 reached abnormality considerably later than the rest of Aβ42/Aβ40 assays across timelines. This finding aligns with and might explain the lower association with amyloid PET observed in our initial cross-sectional study^15^. In addition, the temporal dynamics of plasma biomarkers varied by assay, particularly for Aβ42/Aβ40 in the tau and symptom timelines. These observations may reflect variability in the sensitivity of different types of assays (*e.g.,* mass-spectrometry-based or immunoassays)^7,15,41^. Differences across assays, along with differences in biomarker dynamics, are all important factors to be considered when selecting them for specific uses in trials and clinical practice.

Several previous studies have evaluated AD biomarker trajectories, including plasma biomarkers, using amyloid PET^16,42,43^ or other models^31,44,45^ as proxies of disease progression. However, to the best of our knowledge, only one used a time scale derived from amyloid PET^18^. Of note, the recent study by Li et al.^18^ used an earlier threshold for amyloid positivity, corresponding to the "tipping point” after which individuals accumulated amyloid at a relatively consistent rate (referred to as amyloid onset). Herein, amyloid or tau time were conceptualized as time since a positive scan, as the positivity threshold is consistently used in clinical trials and research studies to define meaningful amyloid accumulation, and it is more directly related to clinically relevant outcomes. Nevertheless, the ordering of biomarker abnormality throughout AD progression was similar in both studies. The current study validated the amyloid PET clock approach in a larger multicenter sample with extensive longitudinal data from individuals at different disease stages and expanded the results with the inclusion of tau PET imaging in addition to key plasma biomarkers for AD measured with different assays, enabling cross-assay comparisons and adding robustness to our findings.

Importantly, we found that the estimated age at tau positivity based on a tau PET clock was a much better predictor of symptom onset than the age at amyloid positivity (R^2^ = 0.86 *vs.* R^2^= 0.38). This result was not unexpected given the extensive evidence supporting that tau is more strongly associated with cognitive decline than amyloid alone^46–49^. However, previous studies had found amyloid onset age to be highly predictive of symptom onset^19,20^. Several factors might be accountable for this difference. Given that ADNI is a multicenter study, there is inherent heterogeneity in clinical diagnosis practices across different sites. Additionally, herein we included participants at all stages of the AD continuum. Still, the predictive value of age at amyloid positivity in our model (R^2^= 0.38) was very close to that of the parental age at symptom onset shown in autosomal-dominant Alzheimer’s disease (ADAD) studies (R^2^ ranging from 0.38 – to 0.56)^50,51^. This is especially noteworthy given that, unlike the expected pure AD pathology in ADAD cases, many factors including comorbid neuropathologies significantly contribute to variability in cognitive symptoms in sporadic AD.

This study has several limitations. First, amyloid PET in ADNI is measured with FBP, which may provide a less sensitive and precise measure of amyloid burden and therefore reduced accuracy of amyloid time estimates compared to previous studies using PIB^19,52^. Second, although the effect of the main AD risk factors such as *APOE* ε4 status or sex on the age at symptom onset was evaluated, there are multiple other variables that could influence the age at symptom onset and were not examined herein (e.g. comorbidities, social determinants of health, genetic variants). Other limitations include a lower sample size for the plasma biomarker analyses compared to the non-plasma biomarkers and cognitive measures, making effects of comparable sizes less likely to be observed among plasma biomarkers. Furthermore, there is a low frequency of dementia cases in our sample, precluding the study of biomarker trajectories in advanced disease stages. Finally, although efforts are being made to increase diversity in ADNI^53^, the present study focused on retrospective data and most participants were white and highly educated, underscoring the need for future studies in more diverse populations.

In summary, we evaluated the timing of change in key AD plasma biomarkers across AD progression timelines using amyloid and tau PET clocks, the latter being highly predictive of symptom onset. Plasma Aβ42/Aβ40 was the earliest to reach abnormal levels consistently across the estimated AD progression timelines, but its dynamic range was limited. In contrast, p-tau217, p-tau181, GFAP and NfL showed utility for disease staging or monitoring. Additionally, our results indicate that plasma biomarker dynamics can vary depending on the timeline and the assay used, which should be considered for their use and interpretation. Importantly, subsequent studies will address the associations of rates of change in plasma biomarkers across AD progression and changes in neurodegeneration and cognitive outcomes, as well as their consistency across assays. Overall, the results of the current study contribute to optimizing the use of plasma biomarkers in clinical trials and clinical practice.

## Data Availability

All data produced in the present study are available upon reasonable request to the authors

## Acknowledgements

The results of the study represent results of the Foundation for the National Institutes of Health (FNIH) Biomarkers Consortium “Biomarkers Consortium, Plasma Aβ and Phosphorylated Tau as Predictors of Amyloid and Tau Positivity in Alzheimer’s Disease” project. The study was made possible through the scientific and financial support of industry, academic, patient advocacy, and governmental partners. We are grateful for the contributions of the following project team members: Anthony Bannon (AbbVie), Michael Baratta (Takeda), Janaky Coomaraswamy (Takeda), Jeff Dage (Indiana University), Iwona Dobler (Takeda), Lei Du-Cuny (AbbVie), Kyle Ferber (Biogen), John Hsiao (NIA), Hartmuth Kolb (formerly with Johnson and Johnson Innovative Medicine), Emily Meyers (Alzheimer’s Association), Yulia Mordashova (AbbVie), William Potter, Maria Quinton (AbbVie), Dave Raunig (Takeda), Erin Rosenbaugh (FNIH), Carrie Rubel (Biogen), Ziad Saad (Johnson and Johnson Innovative Medicine), Patricia Saletti (Alzheimer’s Drug Discovery Foundation), Suzanne Schindler (Washington University in St. Louis), Leslie Shaw (University of Pennsylvania), Gallen Triana-Baltzer (Johnson and Johnson Innovative Medicine), Christopher Weber (Alzheimer’s Association), Henrik Zetterberg (University of Gothenburg). Funding partners of the project include AbbVie Inc., Alzheimer’s Association®, Diagnostics Accelerator at the Alzheimer’s Drug Discovery Foundation, Biogen, Janssen Research & Development, LLC, and Takeda Pharmaceutical Company Limited. Private-sector funding for the study was managed by the Foundation for the National Institutes of Health.

We recognize C2N Diagnostics, Fujirebio Diagnostics with the Indiana University National Centralized Repository for Alzheimer’s Disease and Related Dementias Biomarker Assay Laboratory (NCRAD-BAL), Quanterix, and Roche Diagnostics with the University of Gothenburg for performing the plasma biomarker analysis in this study. The NCRAD-BAL is supported by a cooperative agreement grant (U24 AG021886) awarded to NCRAD by the National Institute on Aging. Elecsys β-amyloid (1–42) CSF, Elecsys Phospho-Tau (181P) CSF and Elecsys Total-Tau CSF assays are approved for clinical use. COBAS and ELECSYS are trademarks of Roche. All other product names and trademarks are the property of their respective owners. The NeuroToolKit is a panel of exploratory prototype assays designed to robustly evaluate biomarkers associated with key pathologic events characteristic of AD and other neurological disorders, used for research purposes only and not approved for clinical use (Roche Diagnostics International Ltd, Rotkreuz, Switzerland).

Finally, we would like to acknowledge ADNI for the plasma samples and ADNI participant data analyzed in this study. Data collection and sharing for the Alzheimer’s Disease Neuroimaging Initiative (ADNI) is funded by the National Institute on Aging (National Institutes of Health Grant U19 AG024904). The grantee organization is the Northern California Institute for Research and Education. In the past, ADNI has also received funding from the National Institute of Biomedical Imaging and Bioengineering, the Canadian Institutes of Health Research, and private sector contributions through the Foundation for the National Institutes of Health (FNIH) including generous contributions from the following: AbbVie, Alzheimer’s Association; Alzheimer’s Drug Discovery Foundation; Araclon Biotech; BioClinica, Inc.; Biogen; Bristol-Myers Squibb Company; CereSpir, Inc.; Cogstate; Eisai Inc.; Elan Pharmaceuticals, Inc.; Eli Lilly and Company; EuroImmun; F. Hoffmann-La Roche Ltd and its affiliated company Genentech, Inc.; Fujirebio; GE Healthcare; IXICO Ltd.; Janssen Alzheimer Immunotherapy Research & Development, LLC.; Johnson & Johnson Pharmaceutical Research & Development LLC.; Lumosity; Lundbeck; Merck & Co., Inc.; Meso Scale Diagnostics, LLC.; NeuroRx Research; Neurotrack Technologies; Novartis Pharmaceuticals Corporation; Pfizer Inc.; Piramal Imaging; Servier; Takeda Pharmaceutical Company Limited; and Transition Therapeutics.

## ADNI Collaborators

Data used in the preparation of this article were obtained from the ADNI database (adni.loni.usc.edu). As such, the investigators within the ADNI contributed to the design and implementation of ADNI and/or provided data but did not participate in the analysis or writing of this report. A complete listing of ADNI investigators can be found at: http://adni.loni.usc.edu/wp-content/uploads/how_to_apply/ADNI_Acknowledgement_List.pdf

## Supplementary material

Supplementary material is available at *Brain* online.

## Appendix 1

This section is included if the article contains an appendix, for example, to list consortium collaborators who must be indexed online.

